# PROTOCOL OF A SYSTEMATIC REVIEW AND METAANALYSIS OF CASE-SERIES AND CASE-REPORTS INVOLVING GLUCAGON IN BETA-BLOCKER OVERDOSE

**DOI:** 10.1101/2023.06.22.23291720

**Authors:** Ramón Aranda Domene, Alberto Esteban-Blanes

## Abstract

The utility of glucagon in human myocardium remains controversial. In the past, some articles have reported inotropic and chronotropic effects of glucagon on the heart, but other studies have found no significant effects. The main cardiovascular indication for glucagon is beta-blocker toxicity (BB), which is listed as recommendation IIb in some guidelines on the basis of some experimental studies and case reports. To our knowledge, no systematic review has been published analyzing the outcomes of glucagon in reports of overdose. This study will attempt to incorporate findings from case series and reports to fill the gap on a topic where experimental and comparative studies have already been conducted. A protocol to the development of this work is provided.

## Introduction

Glucagon is a pleiomorphic hormone that has classically been associated with improvement in cardiac function. Glucagon is thought to have inotropic and chronotropic effects on the myocardium, which has led to its inclusion in the treatment of beta-blocker overdose in some clinical guidelines (1-3). However, controversial reports can also be found in the literature(4-6).

In recent years, two interesting reviews have been published. Rotella et al (7), performed a systematic review evaluating the treatment of beta-blocker poisoning. They concluded that glucagon leads to little improvement in hemodynamics. On the other hand, Petersen et al, studied the hemodynamic effects of glucagon (8). They performed a review that included “in vivo” and “in vitro” studies in animals and human patients-mainly patients with heart failure-and concluded that the weight of evidence of the hemodynamic effects of glucagon was low and that the observations in the published studies were inconsistent. After the review by St. Onge(9), who advised against the use of glucagon in calcium antagonists overdose, beta-blocker toxicity remains the main indication for the use of glucagon in cardiovascular disease.

Recently, some controversial results have emerged in the medical literature. Senetra et al. published a case series of 144 administrations of glucagon in the context of beta-blocker overdose, and the clinical efficacy was low (10). On the other hand, Petersen et al conducted a randomized cross-over trial comparing glucagon with fluids in 10 healthy men who had previously received low doses of esmolol and showed improvements in systolic blood pressure and heart rate after glucagon administration (11).

Recently, our group found no effect of glucagon on human myocardium “in vitro” Furthermore, no glucagon receptors were found by our group (12), suggesting that glucagon has no effect on human myocardium. Nevertheless, intravenous glucagon is considered a Level IIb treatment for beta-blocker poisoning in the 2018 ACC / AHA / HRS Guideline on the Evaluation and Management of Patients with Bradycardia and Cardiac Conduction Delay and the 2020 and 2021 US and European Guidelines on Cardiopulmonary Resuscitation (1-3).

To our knowledge, no other work has been published examining the outcomes of glucagon in reports of beta-blocker poisoning. In addition, Rotella et al included only 9 case reports and 2 case series, and Petersen did not include any study of beta-blocker poisoning, although case reports of glucagon use in beta-blocker overdose are common and often listed in guidelines, leading to the clinical use of glucagon.

In our opinion, an analysis of this underlooked part of the available literature could provide useful information to fill the gap in current knowledge about this hormone and its cardiac effects.

## Objetives

The purpose of this review will be to attempt to answer the the following questions:

- Could glucagon have an effect on hemodynamics or heart rate in patients with beta-blocker overdose according to the case series and report literature?
- Does the literature support the recommendation of some clinical guidelines to use glucagon in beta-blocker overdose?
- Could outcomes in this setting be affected by concomitant administration of other vasoactive agents?

## Methods

A systematic review is performed using the main databases (EMBASE, PUBMED, Google Scholar, WOS, Scholar Semantics, and Cochrane) searching all relevant case series and case reports published before July 2023 on the use of glucagon in beta-blocker overdose. In addition, article references were reviewed to include all available documents related to this work. Unpublished studies are not sought.

This study is conducted according to the recommendations of the Preferred Reporting Items for Systematic reviews and Meta-Analyzes (PRISMA) (13). The protocol was conducted according to the PRISMA P assessment (14).

Two independent reviewers (RA and AE) performed the screening and selection of articles. More detailed information on article selection is presented in the PRISMA diagram. There is no provision for contacting the authors of the reports.

### Search terms

We performed the search using the following terms on Pubmed: (“glucagon” AND “beta blocker” OR “beta antagonist” AND (“intoxication OR overdose OR toxicity OR overdose OR poisoning OR error) NOT (glucagon-like OR GLP).

### Data extraction and quality assessment

Screening and selection of studies and their inclusion in the systematic review are performed by two independent reviewers. No information about the selection will be exchanged between the reviewers. Once the selection of articles is complete, the data are merged and duplicates are removed. Discrepancies are resolved through the intervention of a third author. Reports are recorded in a separate Microsoft Excel spreadsheet until item selection is complete.

The following data is extracted: Author, year of publication, type of article, age, sex, concomitant diseases, type of BB, dose of BB, use of other medications, clinical status before and after intervention, systolic blood pressure or main blood pressure before and after intervention, heart rate before and after intervention, rhythm before and after intervention, dose and route of administration of glucagon, use of other treatments (including fluids) before, with, or after glucagon administration, and mortality. Data extraction is performed by two independent reviewers. RA extracts the data and AE verifies the accuracy of the extracted data. Discrepancies are resolved with the help of a third author (JH). Reports are recorded in a shared Microsoft Excel spreadsheet.

### Quality assessment

Quality assessment of articles is performed as described by Nambiema et al. (15) in their protocol for “Reports and Case Series in Systematic Reviews for Clinical Toxicology,” based on the Navigation Guide instructions.

The assessment is performed at the study level, evaluating the scope of the report, the information presented (dosage and timing of glucagon administration, concomitant treatment, hemodynamic outcomes, etc.), and the context and financial bias of the reports. Two reviewers are involved in the quality assessment, and disagreements between assessments are resolved by a third author. The strength of the evidence is assessed (at GRADE).

### Exclusion criteria

Only articles that include information on glucagon administration and a description of medical status before and after treatment will be considered. There are no restrictions on year of publication, age, or sex. Only case reports and case series in Spanish, French, Portuguese, Italian, and English were considered. Exclusion criteria include animal models, experimental studies, incomplete reports, conference abstracts, randomized trials, and article reviews.

We will only consider case series and case reports. The single RCT (11) evaluating the cardiovascular efficacy of glucagon will be discussed separately. Beta-blocker toxicity with glucagon treatment is not as common, and we estimate that there are no more than 100 references in our review. For this reason, and because Petersen conducted a comprehensive review of observational/experimental articles in 2018, we believe this article may complete the available information on the drug in a cardiovascular setting.

### Definitions

Efficacy was defined in our study as:

- Efficacy alone or no adjuvant efficacy was defined as a combined result of: HR Increase > 10ppm (OR) SBP increase > 20 mmHg (OR) MAP increase > 10 mmHg (OR) clinical improvement after administration of glucagon (bolus or perfusion) before administration of another drug
- Adjuvant efficacy was defined as a combined result of: HR Increase > 10ppm (OR) SBP increase > 20 mmHg (OR) MAP increase > 10 mmHg (OR) clinical improvement after glucagon (bolus or perfusion), even if other vasoactive, inotropic, or chronotropic drugs were administered concomitantly with glucagon.
- No efficacy was defined if the other criteria were not met.

### Statistical analysis

Patients were be divided into three groups according to the results after glucagon administration: Not effective, effective with adjuvant treatment, or effective alone. Statistical analysis by group was performed using chi-square, ANOVA, or Kruskal Wallis as needed. Results were expressed as mean (SD) or median (IQR) according to the normality distribution of each variable.

Subgroup analysis. Metaregression using binary logistic regression. Subgroups included date of publication (before and after 1990), sex, age groups (< 30; 30-60 and > 60 years), major comorbidities (heart failure, chronic kidney disease), high glucagon dosage, concomitant treatment (insulin, ILE, vasoactive drugs…), clinical status, and poisoing with more than two drugs. Tests were performed using SPSS® v.24.0 (IBM Corp. Released 2016. IBM SPSS Statistics for MacOS, version 24.0. Armonk, NY: IBM Corp.).

On the other hand, we will perform a proportional meta-analysis with case series and pooled case reports in which glucagon was used in the treatment of beta-blocker overdose, using Rstudio® v.2022.12.0+353 (16) with the Meta (Metaprop) package (17). Proportional or prevalence meta-analysis has been proposed by some authors as a tool to study and summarize the effectiveness of a treatment for a specific and rare condition (18, 19). A random-effects model (18, 20) using the Freeman-Tukey Double Arscine Transformation (21) of the efficacy of glucagon on systolic blood pressure (SBP) and heart rate (HR) in each article. The results of the fixed-effects models could be also included in the text and figures. Results will be presented as the effectiveness of glucagon in case series and case reports (pooled) with the 95% confidence intervals (CI) based on the weighting of each report. I^2^ and Tau^2^ index were included in the results, although the accuracy of these heterogeneity measures has not been validated in proportional meta-analyzes (18, 20).

Case-series and the pooled case reports will be included in the analysis, and only publications in which glucagon was administered alone were included in the main analysis. Forest plots including cases with adjuvant glucagon efficacy are available in the supplemental materials (Supplemental Materials). Funnel plots were available only in the supplemental materials because of previously reported inaccuracy in detecting publication bias in proportional meta-analyzes (22).

## Data Availability

All data produced in the present study are available upon reasonable request to the authors

## OUTCOMES

We resumed a list of the future variables:

### Baselines and clinical onset

Number of articles

Number of patients

Age

Comorbilities

Sex

Clinical satus at glucagon administration

Type of beta-blocker

Two or more drugs poisoning

Glucagon dosage

Glucagon infusion

Dosage according to clinical guidelines

Effectivity according preset definitions

HR modification

SBP modification

**TABLE 1:** Quality assesment

| *Item* | *Author/Year* | *Rate* | **Rationale for your judgment** |
| --- | --- | --- | --- |
| **DOWNGRADE CATEGORIES** |  |  |  |
| **Rating for Downgrade categories (Study Limitations):**<br>0 no change<br>-1 decrease quality 1 level<br>-2 decrease quality 2 levels |  |  |  |
| **Category 1. Risk of Bias** |  |  |  |
| **Subcategories:** |  |  |  |
| *1.1. Are the study groups at risk of not representing their source populations in a manner that might introduce selection bias?* |  |  |  |
| *1.2. Were exposure/ Intervention (toxic, treatment) assessment methods lacking accuracy?*<br>*Consider:*<br>1) *Identification of the exposure*<br>2) *Dose evaluation*<br>3) *Toxicological values*<br>4) *Clincial effects*<br>5) *Biological effects*<br>6) *Treatments given (dose, timing, route)* |  |  |  |
| *1.3. Were outcome assessment methods lacking accuracy?* |  |  |  |
| *1.4. Was potential confounding inadequately incorporated?* |  |  |  |
| *1.5. Were incomplete outcome data inadequately addressed?* |  |  |  |
| *1.6. Does the study report appear to have selective outcome reporting?* |  |  |  |
| 1.7. *Did the study receive any support from a company, study author, or other entity having a financial interest?* |  |  |  |
| 1.8. *Did the study appear to have other problems that could put it at a risk of bias?* |  |  |  |
| **Category 2. Indirectness of Evidence** |  |  |  |
| **Category 3. Inconsistency of Evidence** |  |  |  |
| **Category 4. Imprecision of Evidence** |  |  |  |
| **Category 5. Publication Bias** |  |  |  |
| **Category 5. Publication Bias** |  |  |  |
| **UPGRADE CATEGORIES** |  |  |  |
| **Rating for Upgrade categories:**<br>0 no change<br>+1 increase quality 1 level<br>+2 increase quality 2 levels |  |  |  |
| **Category 6. Large Magnitude of Effect** |  |  |  |

